# Association between food insecurity, ethnicity, and mental health in the UK: An analysis of the Family Resource Survey

**DOI:** 10.1101/2023.05.26.23290572

**Authors:** Maddy Power, Tiffany Yang, Katie Pybus

**Affiliations:** Department of Health Sciences, University of York; Bradford Institute for Health Research; Hull York Medical School, University of York

**Keywords:** Food insecurity, food poverty, mental health, ethnicity, Family Resource Survey

## Abstract

**Background:** Food insecurity is associated with mental ill-health, but there is limited evidence on ethnicity despite indication that minority ethnic groups are at risk of food insecurity and worse mental health. We assess the relationship between UK food insecurity, ethnicity and mental health using a representative household survey.

**Methods:** Data from the 2019/20 Family Resource Survey provided information on ethnicity, subjective rating of anxiety (10-point scale), presence of long-standing illnesses affecting mental health, and food security assessed using 10-item Adult Food Security module. Linear and logistic regression was used to assess the relationship between food security status and degree of anxiety and presence of long-standing illness affecting mental health. Analyses were adjusted for covariates and stratified by ethnicity.

**Results:** 19,210 participants were included. The majority were food secure (87%), identified as White (90.7%), reported a median and interquartile range of anxiety of 2 (0-5), and 22% reported a long-standing illness affecting mental health. Food insecurity was associated with increased levels of anxiety (adjusted β=1.51, 95% CI:(1.34, 1.68)) among all ethnic groups, with greatest increase among people identifying as Black/African/Caribbean/Black British (β=1.75 (1.05, 2.44)). Food insecurity was associated with longstanding illness affecting mental health (adjusted OR 2.01 (1.70, 2.39)) among all ethnic groups; Asian/Asian British respondents reported the highest odds of having a longstanding illness affecting their mental health (OR=2.63 (1.05, 6.56)).

**Conclusion:** The impact of UK food insecurity on mental health affects all ethnic groups but is worse for ethnic minorities, necessitating a population-wide response to food insecurity alongside targeted interventions addressing ethnic inequalities.

## INTRODUCTION

Household food insecurity is a cause and consequence of poor mental health, as evidenced by an extensive body of North American literature (1, 2). Research posits that the mental health consequences of food insecurity may be numerous and diverse, including chronic depression, anxiety, and feelings of powerlessness (3), while studies suggest that household food insecurity can place a significant additional burden on healthcare services (4).

Over the last decade, food insecurity has become increasingly relevant to the health of United Kingdom (UK) populations due to the continuing retreat of the welfare state, growing inequality, and the impact of sustained public sector austerity following the 2007-08 economic recession (5). Emerging evidence on UK food insecurity points towards a relationship with mental health (6) and shows that food bank usage, an indicator of very low food security (7), is particularly prevalent among individuals who report mental health problems (8). The sharp growth since 2010 and current scale of food bank use suggests a growing public health crisis, but one that remains poorly understood and inadequately conceptualised.

Ethnic minority groups, especially Black British and South Asian groups, are at higher risk of poverty and experience worse mental health than the majority white ethnic group (9, 10). As a result of changes to taxes, benefits and public spending between 2010 and 2020, Black British and South Asian households in the lowest fifth of incomes have experienced the largest average drop in living standards (11). This may indicate that they are at higher risk of food insecurity, and associated mental ill-health, than the majority (white British) ethnic group. However, evidence on the relationship between food insecurity and ethnicity in the UK – including the potentially heightened risk of ethnic minority groups to poor mental health associated with food insecurity – remains limited. This is, in part, attributable to the historical absence of routinely collected national-level data on food insecurity, prior to the publication of the 2019-20 Family Resource Survey (FRS) (in 2021); it is arguably also, however, a consequence of the relative neglect of race and ethnicity in research on UK food insecurity (12).

The descriptive statistics in the FRS evidence ethnic inequality in UK food insecurity, specifically very high food insecurity among Black/African/Caribbean/Black British households and comparatively low food insecurity among and White households (13). Analysis of the nationally representative 2016 Food and You survey (14) similarly illustrated differences in food insecurity by ethnicity. The relatively small size of the survey forces the researchers to use crude categories of ethnic group: ‘white’ and ‘Other ethnic group’/ ‘non-white’. Using these categories their analysis suggests that adults who describe themselves as belonging to a ‘non-white’ ethnic group are more likely to report food insecurity than adults identifying as ‘white’. The researchers also find that ethnicity is associated with moderate food insecurity but not with severe food insecurity, suggesting that ethnicity may be a less important factor in food insecurity than other characteristics, such as income, health status and gender. At a local level, however, the picture may be more complex. Analysis of food insecurity in the Born in Bradford survey (including pregnant women living in the Bradford District) found that Pakistani British women were less likely than white British women to report food insecurity (15). Assessment of food bank usage by ethnic group has also identified an under-representation of ethnic minorities (16, 17).

There is an urgent need for additional research to better establish the demography of food insecurity and mental health in the UK. The development of appropriate targeted health interventions and policy responses to food insecurity is contingent upon this knowledge. Against this context, the study has the following aims:

- to further assess the relationship between food insecurity and ethnicity in the UK;
- to explore the relationship between food insecurity and mental health and assess how this relationship is affected by ethnicity.

## METHODS

### Study population

The Family Resource Survey (FRS) is a continuous household survey which collects information on a representative sample of private households in the UK. Its primary purpose is to provide information to inform the development, monitoring and evaluation of social welfare policy. It provides annual statistics and commentary on circumstances and income from all sources; housing tenure; caring needs and responsibilities; disability (including physical and mental health); pension participation; savings and investment; and self-employment. The 2019-20 FRS (published in 2021) included data for the first time on household food insecurity.

Households in the FRS are defined as one person living alone or a group of people who may not necessarily be related living at the same address and who share cooking facilities and a living space, such as a sitting, dining, or living room. Within each household is a “Household Reference Person” (HRP), who is defined as the highest income householder, and households may include one or more “benefit units” (families); the head of the benefit unit may not necessarily be the HRP.

### Sociodemographic characteristics

Age of the participant was collected in 10-year age range categories from age 16 through age 85 and over. Ethnicity was collapsed from five categories (White; Mixed/multiple; Asian/Asian British; Black/African/Caribbean/Black British; Other ethnic group) into four categories (White; Asian/Asian British; Black/African/Caribbean/Black British; Mixed/multiple/other) to increase sample size for regression models. Cohabitation was defined as “married/cohabitation” if the respondent was married, in a civil partnership, or cohabiting. Participants were coded to “single/divorced/separated/widowed” if they responded they were single, widowed, divorced, had a civil partnership dissolved, or were separated. Housing tenure was categorised to “owned” if the participant owns their home outright or owns with a mortgage, to “rented from council/housing association” if the participant rents from either source, or to “privately rented” if the respondent rents privately, whether furnished or unfurnished.

Household occupancy was determined by summing the number of adults and the number of dependent children within the household. Participants were asked whether they received any state benefits in their own right, including Universal Credit, Housing Benefit, Working Tax Credit, Child Tax Credit, Income Support, Jobseeker’s Allowance, Employment and Support Allowance, Carer’s Allowance, or any/more than one of these; they were categorised as not receiving any benefits if they replied negatively or “none” when asked about each benefit in turn, and categorised as receiving benefits if they responded affirmatively to receiving any benefit.

### Household food security

Household food insecurity was assessed using the 10-item Adult Food Security Survey module. One person was identified by the interviewer as the person with the best information about the food preparation and shopping for the household; this person was chosen to respond to the food security questions for each household. To ensure continuity between responses to the food insecurity questions and other questions in the FRS, food insecurity questions related to a 30-day rather than a 12-month reference period, as used to monitor food insecurity in the US, Canada and worldwide. Analysis suggests that use of the 30-day reference period likely under-estimates annual food insecurity prevalence (Loopstra, nd).

The household food security questionnaire assesses quantitative and qualitative aspects of access to food and food supply and intake, including anxiety or perceived inadequacy of food intake or supply access, and hunger. Each of the questions had affirmative (“yes”, “often true”, “sometimes true”, “3 or more” days) and negative (“no”, “never true”, “2 or fewer” days) responses; each affirmative response was scored a value of 1 and the responses summed across the questions to generate a final score range of 0-10. Households were then categorised into four categories of food security: (1) high food security (score=0); (2) marginal food security (score=1 or 2); (3) low food security (score= 3-5); (4) very low food security (score=6-10). Households with high or marginal food security were considered “food secure” while those with low to very low food security were considered “food insecure”.

### Health outcomes

Participants were queried about their health in the FRS to assess their wider sense of well-being beyond material and financial circumstances. Participants were asked about how anxious they felt the previous day on a scale of 0 for “not at all anxious” to 10 for “completely anxious”. They were additionally asked whether they had any physical or mental health conditions or illnesses lasting or expecting to last for 12 months or more. Responses were coded to “yes” if they responded affirmatively, “no” if they responded negatively or with “don’t know”. Participants who chose not to respond were coded to missing. Among participants who responded affirmatively, they were additionally asked whether any of these conditions affected their mental health (categorised to “yes” or “no”).

### Statistical analysis

All analyses were conducted in R version 4.0.2 (R Foundation for Statistical Computing, Vienna, Austria, 2020). Analyses were conducted for the HRP member of the household (n=19,210). Participant characteristics were described using number (n) and percentage (%) for categorical measures or median and interquartile range (IQR) for continuous measures due to non-Normal data. Characteristics were examined by food security status and their differences assessed using Kruskal-Wallis tests. We used unadjusted and adjusted linear regression to assess the relationship between food security status and anxiety. Unadjusted and adjusted logistic regression and estimated marginal means were used to examine the relationship between food security status and longstanding illness affecting mental health. Directed Acyclic Diagrams (DAGs) were drawn to assist in the selection of covariates for inclusion in the adjusted models (Supplementary file 1); confounding variables adjusted for in the models include age, ethnicity, receipt of benefits, cohabitation, income, household occupancy, and housing tenure.

## RESULTS

### Sample characteristics

Table 1 outlines the demographic characteristics of the sample. Overall, there were fewer female than male respondents (41.7% compared to 58.3%). The majority of the sample identified as White (90.7 %); 4.8% reported their ethnicity as Asian/Asian British and 2.5% were from a Black, African, Caribbean or Black British background. Two percent of respondents described themselves as being from a mixed or “other” ethnic background. Around two thirds of the sample (64.6%) owned their home outright or with a mortgage. A fifth (20.1%) of respondents reported being in receipt of social security payments and the mean household income was £610 (£364-£1,032) per week, before housing costs. Less than half responded as having a longstanding illness (44%) and just over a fifth of those respondents (22.3%) reported that their longstanding illness affected their mental health.

**Table 1.** Characteristics of the head of household

|  | **Total**<br>**n=19210** |  |
| --- | --- | --- |
|  | **N** | **Median[IQR]/%** |
| **Sex** |  |  |
| Female | 8066 | 41.7 |
| Male | 11204 | 58.3 |
| **Age** |  |  |
| 16-24 | 451 | 2.3 |
| 25-34 | 2300 | 12 |
| 35-44 | 3109 | 16.2 |
| 45-54 | 3352 | 17.4 |
| 55-59 | 1797 | 9.4 |
| 60-64 | 1689 | 8.8 |
| 65-74 | 3528 | 18.4 |
| 75-84 | 2984 | 15.5 |
| 85+ |  |  |
| **Ethnicity** |  |  |
| White | 17422 | 90.7 |
| Asian/Asian British | 929 | 4.8 |
| Black/African/Caribbean/Black British | 475 | 2.5 |
| Mixed/multiple/other | 384 | 2 |
| **Marital/cohabitation** |  |  |
| Married/civil partnership/cohabitation | 10738 | 55.9 |
| Single/divorced/widowed/separated | 8472 | 44.1 |
| **Tenure** |  |  |
| Owned | 12404 | 64.6 |
| Privately rented | 3262 | 17 |
| Rented from council/housing association | 3544 | 18.4 |
| **Household occupancy** | 19210 | 2 (1, 3) |
| **In receipt of benefits** |  |  |
| No | 15353 | 79.9 |
| Yes | 3857 | 20.1 |
| **Household income** | 19210 | 610 (364, 1032) |
| **Food security** |  |  |
| High | 16651 | 86.7 |
| Marginal | 1069 | 5.6 |
| Low | 741 | 3.9 |
| Very low | 749 | 3.9 |
| **Longstanding illness affecting mental health** |  |  |
| No | 6593 | 77.7 |
| Yes | 1890 | 22.3 |
| **Anxiety** | 16729 | 2 (0, 5) |

### Demographic characteristics by food security status

We observed differences across all demographic characteristics by food security status with the exception of household occupancy (Table 2).

**Table 2.** Distribution of characteristics by food security status

|  | Food insecure<br>n=1490 (7.8%) |  | Food secure<br>n=17720 (92.2%) |  | p-value* |
| --- | --- | --- | --- | --- | --- |
|  | N | Median[IQR]/% | N | Median[IQR]/% |  |
| **Sex** |  |  |  |  | <0.001 |
| Female | 916 | 61.5 | 7090 | 40 |  |
| Male | 574 | 38.5 | 10630 | 60 |  |
| **Age** |  |  |  |  | <0.001 |
| 16-24 | 73 | 4.9 | 378 | 2.1 |  |
| 25-34 | 286 | 19.2 | 2014 | 11.4 |  |
| 35-44 | 378 | 25.4 | 2731 | 15.4 |  |
| 45-54 | 353 | 23.7 | 2999 | 16.9 |  |
| 55-59 | 149 | 10 | 1648 | 9.3 |  |
| 60-64 | 113 | 7.6 | 1576 | 8.9 |  |
| 65-74 | 105 | 7 | 3423 | 19.3 |  |
| 75-84 | 33 | 2.2 | 2951 | 16.7 |  |
| 85+ |  |  |  |  |  |
| **Ethnicity** |  |  |  |  | <0.001 |
| White | 1257 | 84.4 | 16165 | 91.2 |  |
| Asian/Asian British | 84 | 5.6 | 845 | 4.8 |  |
| Black/African Caribbean/Black British | 95 | 6.4 | 380 | 2.1 |  |
| Mixed/multiple/other | 54 | 3.6 | 330 | 1.9 |  |
| **Cohabitation** |  |  |  |  | <0.001 |
| Married/civil partnership/cohabitation | 392 | 26.3 | 10346 | 58.4 |  |
| Single/divorced/widowed/separated | 1098 | 73.7 | 7374 | 41.6 |  |
| **Tenure** |  |  |  |  | <0.001 |
| Owned | 253 | 17 | 12151 | 68.6 |  |
| Privately rented | 398 | 26.7 | 2864 | 16.2 |  |
| Rented from council/housing association | 839 | 56.3 | 2705 | 15.3 |  |
| **Household occupancy** | 1490 | 2 (1, 3) | 17720 | 2 (1, 3) | 0.6 |
| **In receipt of benefits** |  |  |  |  | <0.001 |
| No | 460 | 30.9 | 14893 | 84 |  |
| Yes | 1030 | 69.1 | 2827 | 16 |  |
| **Household income** | 1490 | 376 (242, 542) | 17720 | 642 (380, 1070) | <0.001 |
| **Long-standing illness affecting mental health** |  |  |  |  | <0.001 |
| No | 393 | 41.7 | 6200 | 82.2 |  |
| Yes | 549 | 58.3 | 1341 | 17.8 |  |
| **Anxiety** | 1388 | 5 (2, 8) | 15341 | 2 (0, 5) | <0.001 |

\*Chi-squared or Kruskal-Wallis test for differences between food security status

Food insecure households were younger, with almost two-thirds aged 25-54 (63.8%), had more participants identifying as non-White (15.6% compared to 8.8%), female (61.5% compared to 40%), and more likely to be single, divorced, widowed, or separated (73.7% compared to 41.6%). Food insecure households were less financially secure compared to those who were food secure, reporting a lower income, being less likely to own their own home, and more likely to be in receipt of any benefits. A higher proportion also reported a longstanding illness which affected their mental health (58.3% compared to 17.8%) and higher levels of anxiety.

### Food security status, ethnicity and mental health

Food insecurity was associated with increased levels of anxiety among all ethnic groups (Table 3). Food insecurity was associated with increased levels of anxiety in both unadjusted (β=2.18 95% Confidence Intervals (CI)): (2.02, 2.33)) and adjusted (β=1.51 (1.34, 1.68) models as well as increased odds of having a longstanding illness affecting mental health (unadjusted: Odds Ratio (OR) 6.46 (5.60, 7.45); adjusted: 2.01 (1.76, 2.39)).

**Table 3.**
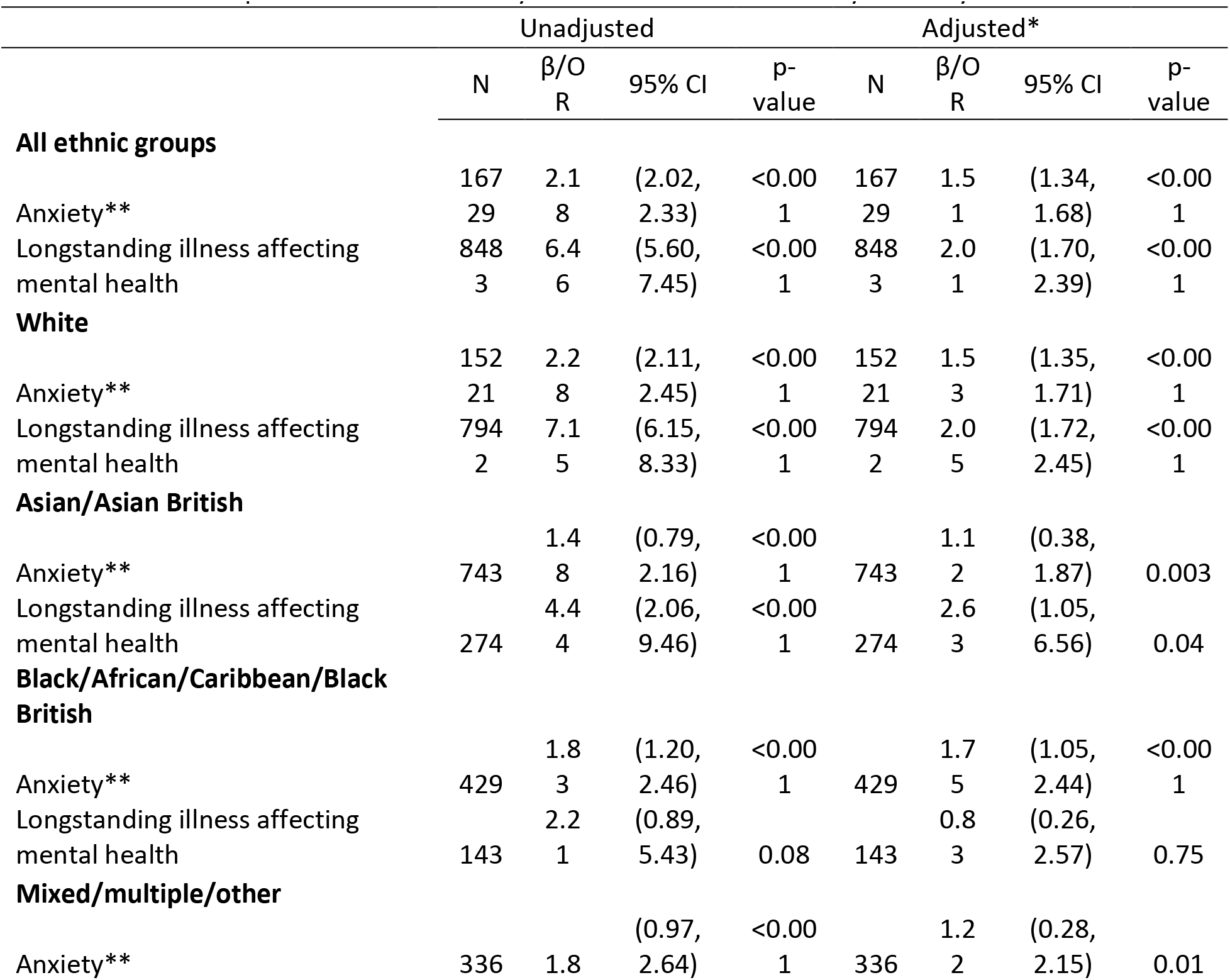
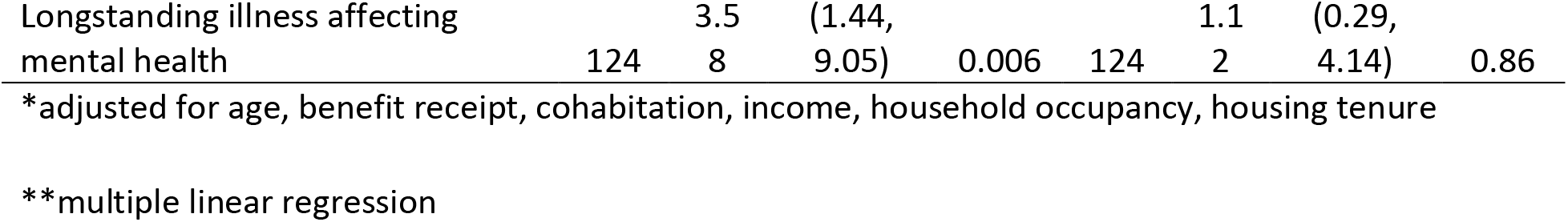
Relationship between food security status and mental health by ethnicity

When adjusted for covariates, food insecure participants identifying as Black/African/Caribbean/Black British had higher anxiety levels (β=1.75 (1.05, 2.44)) than other ethnic groups. Respondents from an Asian/Asian British background reported the highest odds of having a longstanding illness affecting mental health (OR=2.63 (1.05, 6.56)) followed by those identifying as White (OR=2.05 (1.72, 2.45)).

## DISCUSSION

These analyses augment growing evidence on UK food insecurity, identifying demographic differences in food insecurity and in its relationship with mental health. In our study, food insecurity was associated with unstable circumstances such as being younger, single, divorced or widowed, renting, having a lower income, and being in receipt of benefits. Aligning with existing evidence (14), we identified substantial ethnic differences in food insecurity – notably, 20% of Black/African/Caribbean/Black British respondents are food insecure compared to 7% of White British respondents.

We identified a relationship between food insecurity and mental health, specifically between food insecurity and both anxiety and reporting a long-standing health condition affecting mental health (12 months or more). Stratifying by ethnicity, we found that the degree of worse mental health reported is greater for some minority ethnic groups. For example, being food insecure for Black, African, Caribbean, Black British respondents is associated with a larger increase in subjective anxiety than for other ethnic groups reporting food insecurity. In our analyses, the Black, African, Caribbean, Black British group incorporates a higher proportion of females, renting, benefit receipt and lower income, which are all associated with being food insecure; the relationship we identify here between food insecurity and anxiety among Black respondents could reflect a broader composite hardship in which the Black population experiences multiple forms of overlapping disadvantage and discrimination of which food insecurity and anxiety is part.

In contrast, food insecure Asian/Asian British respondents had higher odds of reporting a long-term mental health condition than other ethnic groups reporting food insecurity. Evidence suggests that low income Asian and Asian British households are more likely than other ethnic groups to have access to strong social and familial networks which may mitigate the negative health impacts of moderate food insecurity (18, 19), but also that food insecurity can be highly shameful in Asian and Asian British communities and likely to be underreported (20). There is an established relationship between disability and severe food insecurity (21); it is possible that Asian/Asian British people experiencing severe food insecurity are at high risk of disability and simultaneously are more likely than other ethnic groups to be ostracised from familial and social support, placing them at risk of isolation, financial hardship and worsening mental health.

It is also possible that worse mental health among food insecure ethnic minority respondents is a consequence of exclusion from and challenges, such as racism and stigma, surrounding access to mental health services (22-24) and to food banks (16, 17). Such social exclusion may exacerbate the negative mental health consequences of food insecurity and compound pre-existing hardship.

### Strengths and limitations

This is one of the first studies to explore the demography of food insecurity in a large nationally representative UK sample and the first to consider ethnic differences in the relationship between food insecurity and mental health in the UK population. As such, it makes a significant contribution to the evidence on UK food insecurity, highlighting the complex relationship between food insecurity, ethnicity and health. The study is, nevertheless, subject to limitations. It is possible that our findings are an underestimate of food insecurity. On the advice of the Office for National Statistics (ONS), the analysis was conducted using the Household Reference Person (HRP), who is the householder with the highest income or, where two people in the household have the same income, the older of the two. The use of the HRP in our analyses of food insecurity likely explains the lower number of female respondents (41.7% compared to 51% in the general population (25)) who are more likely to report food insecurity than men (26) and possibly also the lower number of minority ethnic respondents (90.7% of the sample identified as White compared to 81.7% in the general population (25)). Households were considered to be “food insecure” if they reported low to very low food security while households who reported marginal food security were categorised as “food secure”; there is some debate as to whether marginal food security is a reflection of food insecurity rather than food security and categorising the variables in this way misrepresents the realities/scale of food insecurity in the population. The food insecurity questions apply to a 30-day rather than a 12-month reference period, likely further contributing to an underestimation of food insecurity (27). The cross-sectional nature of the data makes it impossible to conclude the direction of the relationship between food insecurity and mental health. Assessing the direction of the relationship is additionally complicated by the variable time periods of the food insecurity and long-standing illness questions: the length of the longstanding illness (12 months) implies that it could potentially start before and hence be a precursor to any food insecurity (30 days). Future research could apply (quasi) experimental approaches to get closer to causal estimates.

## Conclusion and implications

Using data from 2019/20, this study finds a strong relationship between food insecurity and mental health among all ethnic groups. Since this data was collected, there have been sharp increases in both food insecurity and mental illness in the UK, first as a consequence of the Covid-19 pandemic and then as a result of rapid rises in the cost of food, energy and housing (28). Our findings are likely to be an underestimate of current levels of food security and thus the potential consequences for mental health today may be greater than we have been able to ascertain using 2019/20 data. Policy makers should urgently address rising food insecurity among all groups while simultaneously employing targeted and culturally appropriate interventions to tackle ethnic inequalities in both food insecurity and mental health.

## Data Availability

All data in the present study are publicly available at GOV.UK.

https://www.gov.uk/government/collections/family-resources-survey--2

## Competing Interests

None declared

## Funding

The research was funded by a Wellcome Trust Research Fellowship held by the first author (Grant number: 221021/Z/20/Z).

## Ethical Approval

The study uses publicly available ONS secondary data. This data, which is in the public domain, is fully anonymised; access and use of this data poses no ethical risk. The data is available for public use via GOV.UK.

## Notes

### Competing Interest Statement

The authors have declared no competing interest.

