## Supplementary figures and images for "Association between food insecurity, ethnicity, and mental health in the UK: An analysis of the Family Resource Survey"

### Supplementary Figure 1A

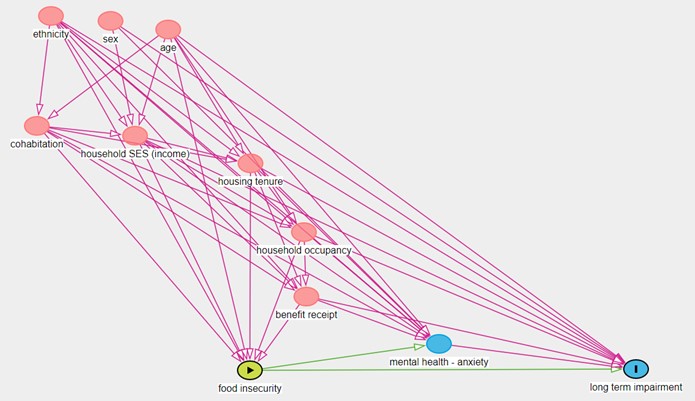

### Supplementary Figure 1B

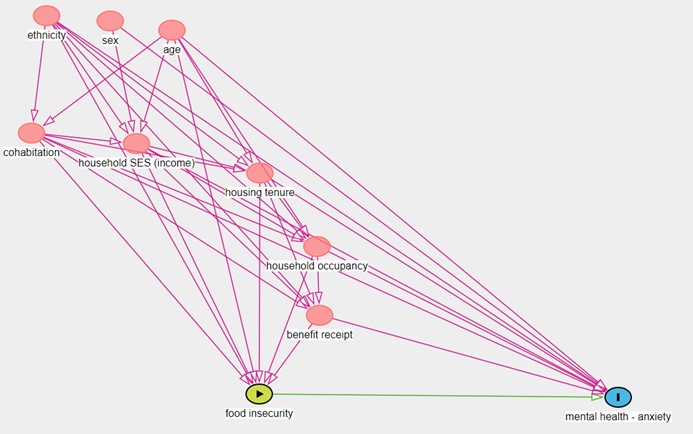
